# State Variation in Neighborhood COVID-19 Burden: Findings from the COVID Neighborhood Project

**DOI:** 10.1101/2023.05.19.23290222

**Authors:** Grace A Noppert, Philippa Clarke, Andrew Hoover, John Kubale, Robert Melendez, Kate Duchowny, Sonia T Hegde

## Abstract

A lack of fine, spatially-resolute case data for the U.S. has prevented the examination of how COVID-19 burden has been distributed across neighborhoods, a known geographic unit of both risk and resilience, and is hampering efforts to identify and mitigate the long-term fallout from COVID-19 in vulnerable communities. Using spatially-referenced data from 21 states at the ZIP code or census tract level, we documented how the distribution of COVID-19 at the neighborhood-level varies significantly within and between states. The median case count per neighborhood (IQR) in Oregon was 3,608 (2,487) per 100,000 population, indicating a more homogenous distribution of COVID-19 burden, whereas in Vermont the median case count per neighborhood (IQR) was 8,142 (11,031) per 100,000. We also found that the association between features of the neighborhood social environment and burden varied in magnitude and direction by state. Our findings underscore the importance of local contexts when addressing the long-term social and economic fallout communities will face from COVID-19.

## Introduction

In the United States (U.S.), the COVID-19 pandemic has affected nearly every American and nearly every part of American life. However, the burden of COVID-19 has not been equally distributed across neighborhoods within the U.S. A growing number of studies shows that COVID-19 has and continues to disproportionately affect neighborhoods comprised of majority racial/ethnic minoritized groups and low-income communities.(*1-13*) Neighborhoods are a source of both risk and resilience to COVID-19 and its long-term sequelae due to their social and economic characteristics (e.g., crowding, housing density, affluence, business types, and political partisanship).(*14-20*) Since the start of the COVID-19 pandemic, structurally disadvantaged neighborhoods (e.g., low SES and/or communities of color) have faced substantially greater population losses, economic hardships, and business closures compared to less disadvantaged neighborhoods, particularly those neighborhoods with a greater share of working-aged adults. For example, in the early months of the pandemic, a 7-state study of COVID-19 prevalence by ZIP code found a higher burden of disease in socioeconomically disadvantaged ZIP codes in Illinois and Maryland.(*1*) A 2021 study using data from the U.S. Household Pulse Survey found that individuals living in states with high levels of pre-pandemic poverty and non-Hispanic Black populations experienced a greater number of COVID-19 hardships including food insufficiency, loss of income, unemployment, and housing instability and that racialized minorities had a slower recovery from these hardships than their White counterparts.(*21*)

Despite initial evidence indicating neighborhoods play an important role in shaping COVID-19 risk and resilience, efforts to fully understand variation across neighborhoods in the U.S. have been hampered by a lack of fine-scale spatially resolute COVID-19 case data. To date, the majority of studies that have examined spatial trends in COVID-19 burden have relied on county-level case data, which prevents examination of local level variation in both the distribution of COVID-19 burden as well as the predictors of COVID-19 burden.(*22-25*) These local patterns are critical to our understanding of which communities will continue to face short-term and long-term health, social and economic consequences from COVID-19. Indeed, a number of studies suggest there is significant heterogeneity in the distribution of COVID-19 within counties and within states.(*8, 26-28*) However, studies that have examined finer spatial levels (e.g., ZIP code and/or census tract) thus far have been limited to a single city, region, or state.(*1-3, 6-12, 28-44*) Some of these studies have examined a group of cities, regions, or states,(*8, 34, 36, 42*) but only a few have had the data to examine patterns across a wider geographic scope.(*1, 29*) To the best of our knowledge, no studies have examined neighborhood-level variation in COVID-19 for more than seven states and none have leveraged existing data sources to explore the influence of neighborhood social conditions in relation to COVID-19 surveillance data at the local-level.

In response to the lack of fine-scale COVID-related spatial data, we launched the COVID Neighborhood Project (CONEP) in 2021. CONEP is a repository of locally-referenced (census tract or ZIP code tabulation area (ZCTA)) COVID-19 case data from April 2020 to April 2022. The repository currently includes cumulative case data for 21 states in all five regions of the U.S. (West, Southwest, Midwest, Northeast, and Southeast). Data collection for the remaining states is ongoing. In this paper, we leverage data from CONEP to illustrate how neighborhood-level COVID-19 burden varies dramatically between states and examine the neighborhood social characteristics that are related to burden. We address a critical gap in the literature by examining the following two research questions: (1) How is COVID-19 distributed across neighborhoods (i.e., ZCTAs or census tracts) within states, and is the neighborhood distribution of COVID-19 burden similar between states? (2) Within each state, is neighborhood COVID-19 burden associated with salient features of the neighborhood social environment?

As previous studies of both COVID-19, as well as many other infectious diseases, have consistently demonstrated the importance of neighborhood socioeconomic status (SES) in determining the distribution of infectious disease burden (*1, 23, 45-47*), we focus on two specific measures of neighborhood SES: neighborhood disadvantage and neighborhood affluence. We also employ measures of neighborhood population density and county-level political partisanship. Given that contact with an infectious pathogen is a necessary cause of infectious disease, we use neighborhood population density as an imperfect proxy of the probability of encountering an infectious case of COVID-19.(*46*) Further, we use a measure of county-level political partisanship to begin to understand the complex role that political ideology has had in shaping COVID-19 testing, vaccination access, and mitigation strategies.(*48, 49*)

## Results

### Cumulative case counts are heterogenous between states

Of the spatially-referenced COVID-19 case data from 21 states, five states had data at the census-tract level and 16 states had data at the ZIP code level (**Table 1**). Case data spanned from April 2020 through April 2022, although there was state variability in the temporal reporting coverage. We calculated the cumulative neighborhood case count (census-tract or ZCTA) per 100,000 population for the entire two-year period and categorized the case counts into deciles for all 21 states (**Figures S1-S3**). Oregon had the lowest cumulative case count of 4,417 cases per 100,000 population and Wisconsin the highest with 31,422 cases per 100,000 population for the two-year period (**Table 2, Figure 1**). Additionally, multiple states had spatial units with population residing in them that were missing case data (see **Table S1**).

**Figure 1.**
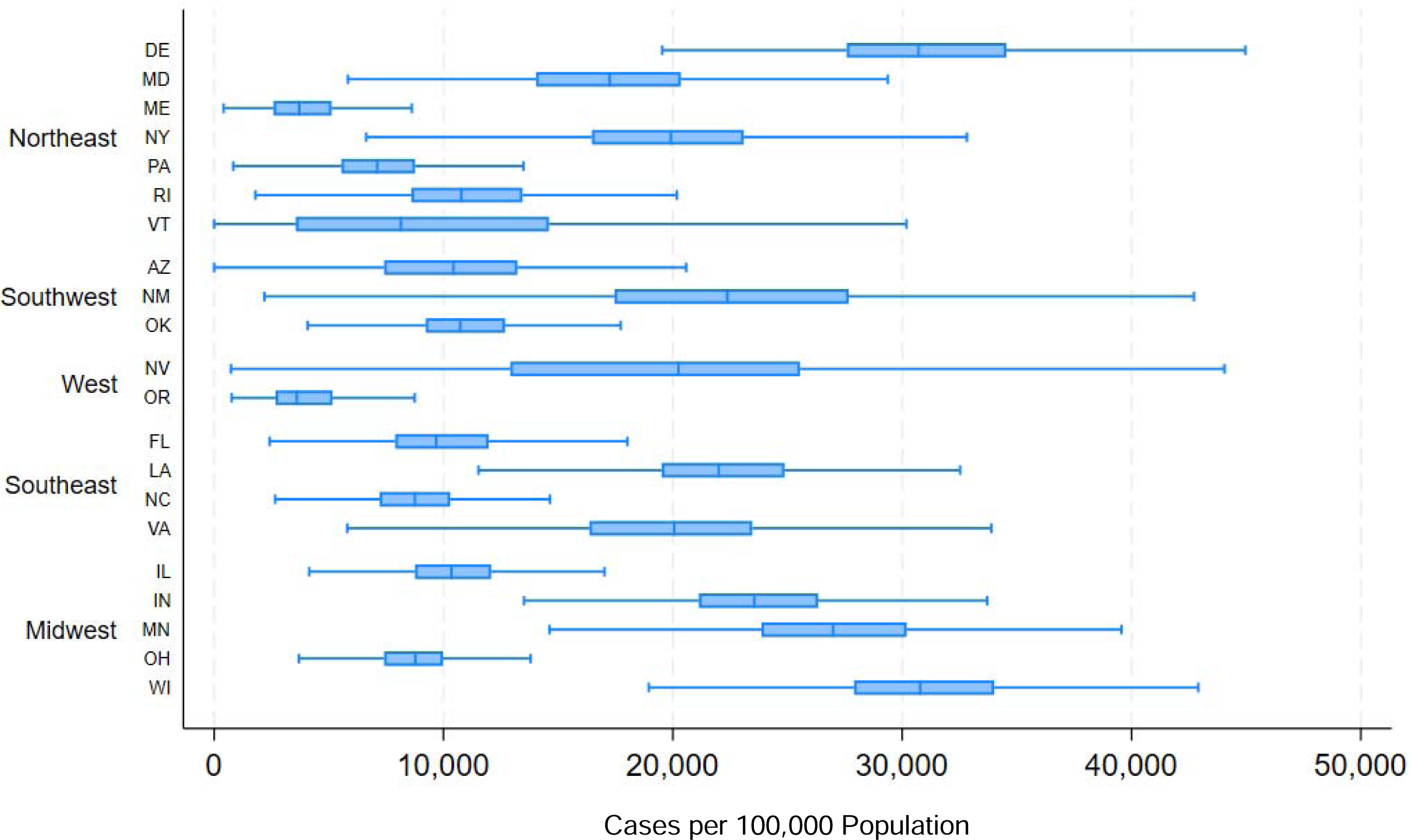
Median and IQR for COVID case counts per 100,000 for each state by region.

**Table 1.**
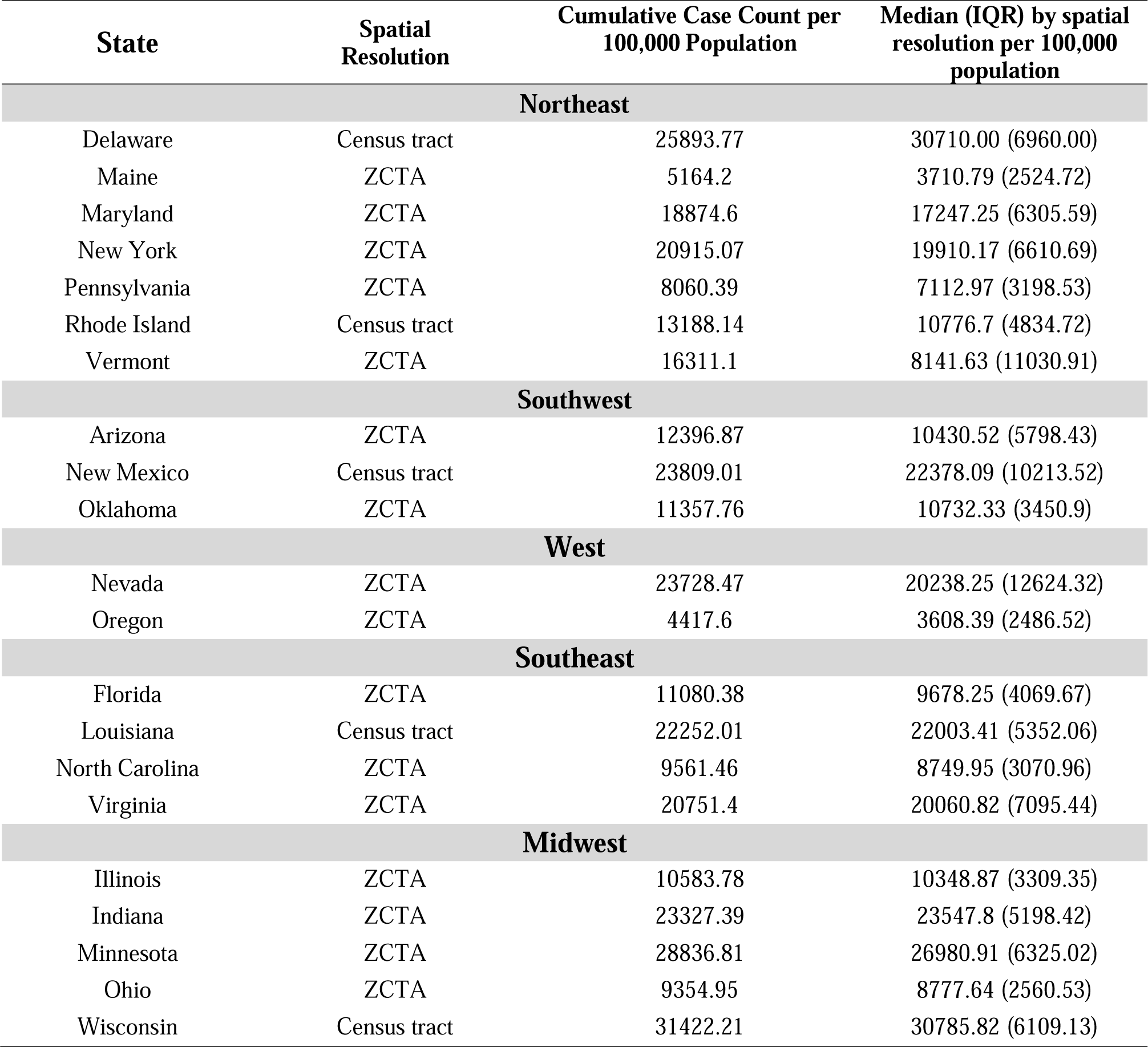
States for which data was obtained, the corresponding spatial resolution, and cumulative COVID-19 case counts per 100,000 population (April 2020-April 2022).

### Neighborhood-level trends in COVID-19 burden differ by state

We calculated the median case count per 100,000 population and interquartile range (IQR) by ZCTA (**Table 3a**) and census tract (**Table 3b**). The median case count per 100,000 population at the neighborhood-level mirrored the trends of the cumulative case count per 100,000. The IQR, however, gives an indication of the neighborhood-level variability in median case counts within each state, and there were notable state-level differences in the IQR of the neighborhood-level cases counts. For example, the highest IQRs were observed in Nevada (IQR = 12,624 per 100,000 population), Vermont (IQR = 11,030 per 100,000 population), and New Mexico (IQR = 10,214 per 100,000 population) which demonstrate a high within-state variability in the distribution of COVID-19 cases at the neighborhood-level. In contrast, the low IQRs for states such as Maine (IQR = 2,525 per 100,000 population), Ohio (IQR = 2,561 per 100,000 population), and North Carolina (IQR = 3,071 per 100,000 population) suggest a more homogenous spread of COVID-19 cases across neighborhood within those respective states.

### Direction and strength of associations between neighborhood social factors and neighborhood COVID-19 burden differ by state

To address the second research question, whether COVID-19 is associated with features of the neighborhood social and physical environment within a state, we used a series of Poisson regression models to estimate the incidence rate ratio (IRR) of case counts per 100,000 population for each neighborhood within each state. We used the recent framework by Noppert, Hegde, and Kubale (2022) to select features of the neighborhood environment that may be particularly relevant for understanding COVID-19 burden, and that may also lend themselves to intervention.(*50*) Their framework posits that infectious disease burden is a function of two primary pathways which may operate at both the individual- and neighborhoods-level: <u>exposure,</u> factors that increase the probability of exposure to an infectious pathogen, and <u>susceptibility</u> factors that increase the likelihood of being infected if exposed. See **Figure 2** for the conceptual diagram used to guide the selection of neighborhood social and physical features.

**Figure 2.**
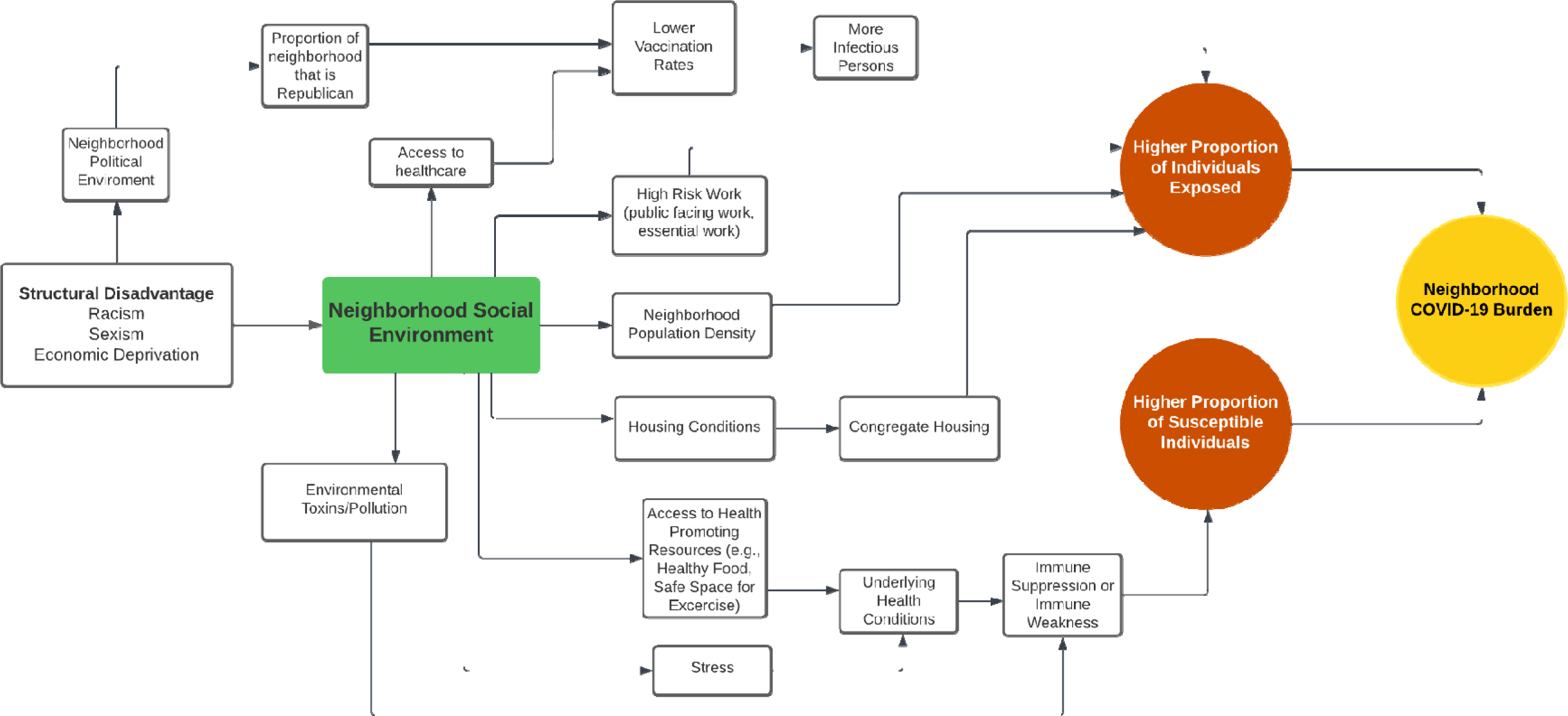
Conceptual diagram describing features of the neighborhood social environment may influence the neighborhood burden of COVID-19.

Based on this conceptual framework, we focused on associations between COVID-19 burden and neighborhood disadvantage and neighborhood affluence with additional control variables for population density and county-level political partisanship. We modeled neighborhood disadvantage and neighborhood affluence as two separate exposures given that they capture two distinct measures of neighborhood SES which is described further in the Methods section below.

We observed associations between neighborhood disadvantage and neighborhood COVID-19 burden for many states (**Figure 3, Table S2**), however, the magnitude and direction of the association differed widely between states. Further, there were no detectable trends by region of the U.S. In 14 states there was a positive association between neighborhood disadvantage and the COVID-19 burden wherein higher quartiles of neighborhood disadvantage (i.e., those more disadvantaged) were associated with a higher IRR of COVID-19. For example, in the highest quartile (Q4) of disadvantaged neighborhoods in Maryland, the IRR was 1.39 (95% CI: 1.21, 1.60) times that of neighborhoods in the lowest quartile of disadvantage (Q1, i.e., those least disadvantaged). Similarly, in Oregon neighborhoods, the highest quartile of disadvantage (Q4) had 1.94 (95% CI: 1.51, 2.51) times the rate of COVID-19 compared to neighborhoods in the lowest quartile of disadvantage (Q1). These associations were robust to additional controls for county-level political affiliation (**Figure 3**).

**Figure 3.**
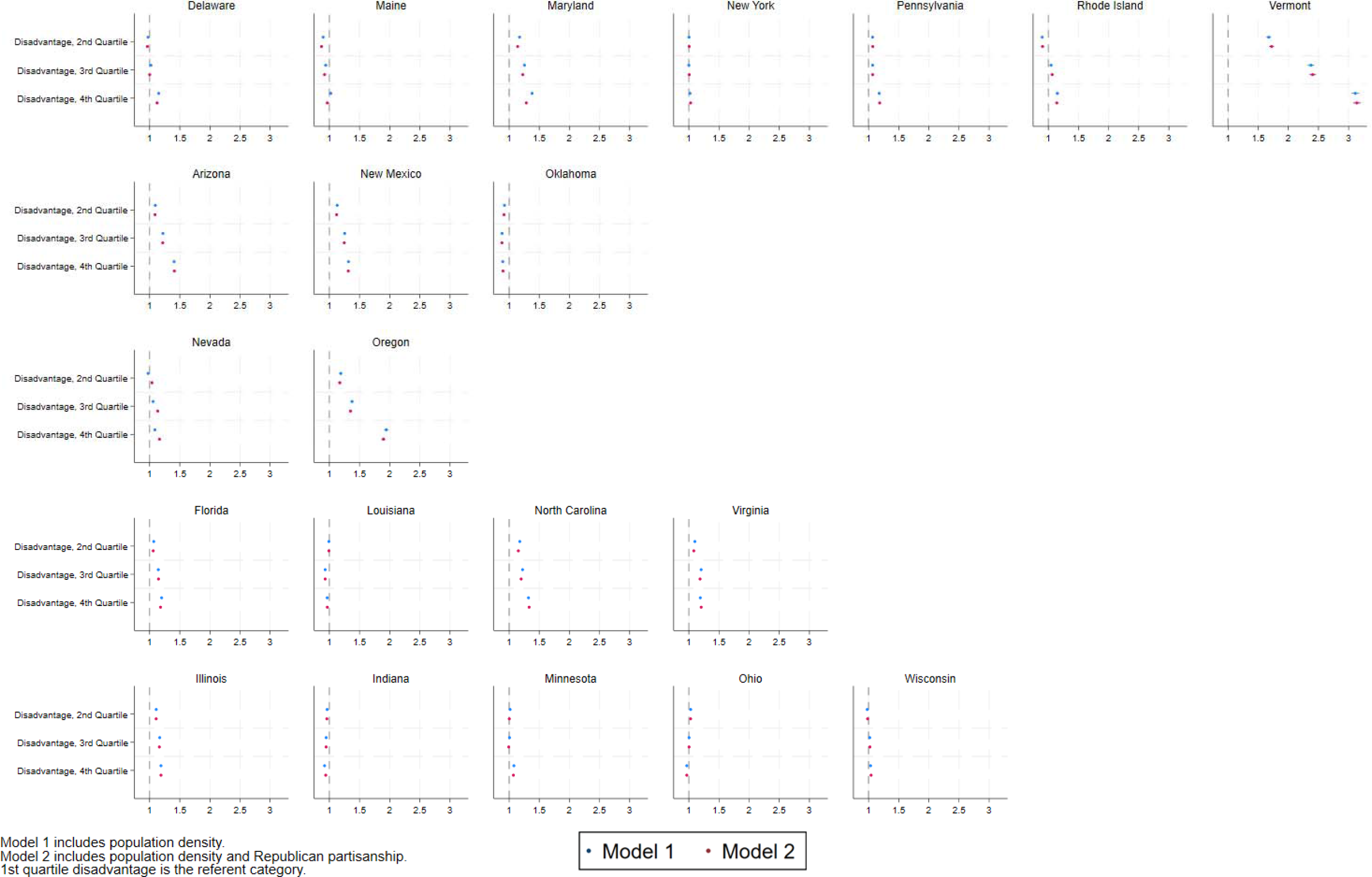
Results of the regression results estimating the association between neighborhood disadvantage and neighborhood COVID-19 burden. Model controls for neighborhood population density. Model 2 adds a control variable for county level political affiliation.

Conversely, in multiple states we observed estimates suggestive of a negative association between neighborhood disadvantage and COVID-19 burden in which neighborhoods in the highest quartile (Q4) of disadvantage had lower rates of COVID-19 compared to states in the lowest quartile (Q1) of disadvantage. However, only in Oklahoma was this association statistically significant. In Oklahoma, neighborhoods in the highest quartile of disadvantage (Q4) had an 11% lower (b = 0.89, 95% CI: 0.81-0.98) incidence rate of COVID-19 compared to neighborhoods in the lowest quartile of disadvantage (Q1). The association was attenuated, though robust, when controlling for county-level political affiliation (**Figure 3**).

We also estimated the association between neighborhood affluence and COVID-19 burden (**Figure 4, Table S3**). Again, we found that the direction and magnitude of the association differed by state with no clear patterns by region. There were 10 states where we observed a negative association between neighborhood affluence and COVID-19 burden. In these states, neighborhoods in the highest quartile of affluence (Q4; i.e., the most affluent) had statistically significant lower COVID-19 burden compared to neighborhoods in the lowest quartile of affluence (Q1). For example, in Arizona neighborhoods in the highest quartile of affluence (Q4), we observed a 25% lower (b=0.75, 95% CI: 0.61, 0.92) rate of COVID-19 compared to neighborhoods in the lowest quartile of affluence (Q1). In Oregon, neighborhoods in the highest quartile of affluence (Q4) had a 48% lower (b=0.52, 95% CI: 0.41, 0.66) rate of COVID-19 compared to neighborhoods in the lowest quartile (Q1). Both associations were robust to controls for county-level political affiliation (**Figure 4**).

**Figure 4.**
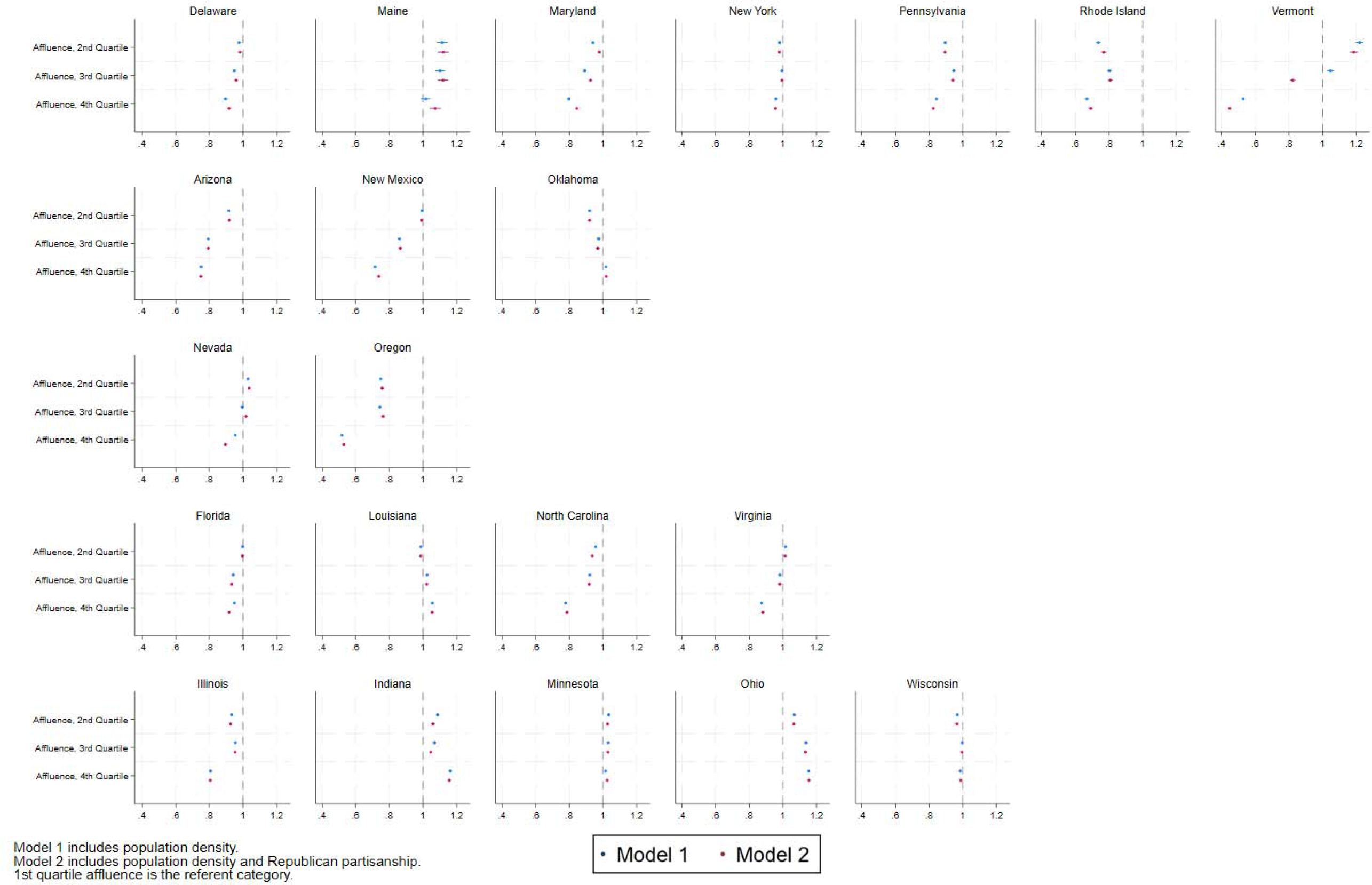
Results of the regression results estimating the association between neighborhood affluence and neighborhood COVID-19 burden. Model controls for neighborhood population density. Model 2 adds a control variable for county level political affiliation.

In contrast, there were multiple states in which we observed estimates suggestive of a positive association between neighborhood affluence and COVID-19 burden. In Indiana, for example, neighborhoods in the highest quartile of affluence (Q4) had an IRR 1.16 (95% CI: 1.07, 1.27) times that of neighborhoods in the lowest quartile of affluence (Q1). The association was robust to controlling for county-level political affiliation (**Figure 4**). In Ohio, neighborhoods in the highest quartile of affluence (Q4) had COVID-19 rates 1.15 (95% CI:1.11, 1.20) times that of neighborhoods in the lowest quartile (Q1) of affluence. The association in Ohio was robust to controls for county-level political affiliation (**Figure 4**).

## Discussion

Our results are the first findings from the COVID Neighborhood Project, a novel data effort designed to collate spatially-referenced COVID-19 data at the neighborhood-level across the U.S. The findings from this study suggest that the distribution of COVID-19 between neighborhoods is dependent on the state context. For some states, there is wide variation in the neighborhood COVID-19 burden (i.e., Nevada, New Mexico, and Vermont), a signal that some neighborhoods account for a disproportionate burden of COVID-19 cases compared to other neighborhoods in the same state. In contrast, for other states, the burden of COVID-19 is more homogenously distributed across neighborhoods (i.e., Maine, Ohio, and North Carolina). These state-level differences may in part be driven by heterogeneity in the neighborhood social environment and local political partisanship or policies, but the exact reasons will take time to elucidate as our results bear out. Though the results from the current investigation are preliminary and observational in nature, they underscore that local neighborhood patterns and dynamics may play an underappreciated role in the distribution and intensity of COVID-19 burden and the long term sequalae that will be endured. These findings hint at the complex relationship between state-and local-based policies, neighborhood features, and the myriad ways in which the interaction of these area-based forces may have influenced the COVID-19 burden felt by millions. Indeed, policy makers should be wary in crafting a one-size-fits-all approach for pandemic mitigation and recovery efforts. Among the states included in this analysis, there is no single, unifying story to describe how COVID-19 has been distributed across neighborhoods within states in the U.S.

There have been numerous studies relating aspects of the social and built neighborhood environment to infectious disease in the U.S.(*47, 51-55*). For many of these studies, a traditional paradigm prevails in which more disadvantaged neighborhoods experience a higher burden of disease (*46, 47, 52*). While our findings support this paradigm for some states, these historically accepted patterns of the relationship between the neighborhood environment and infectious disease burden are challenged in other states; this paradigm is more nuanced and complex than previously imagined. For example, in Oklahoma, less disadvantaged neighborhoods had higher COVID-19 incidence rates even after controlling for political partisanship and population density, a possible artifact of rurality or limited access to health facilities where a case would be recorded. Though our findings are preliminary, in that we are limited by the data we have been able to collect thus far both in geographic scope (i.e., not all 50 states) and other key confounding variables (e.g., neighborhood vaccination rates, access to vaccination sites, access to health facilities, and test positivity rates at the local area level), these broad signals offer a window into the complicated relationship that exists between aspects of the social and built neighborhood environment, state and local policy, and COVID-19 burden. As shown in other studies, there are state-specific behaviors and policies that have shaped the variation in the COVID-19 burden across the U.S. (*49, 56-59*). However, in this study, we demonstrate how the collection of localized data can expand on these results to decompose how state-specific behaviors interact with local area-level concentrated disadvantage or affluence in ways that we are only beginning to gain insight into.

For instance, in one of the few studies that focused exclusively on understanding the impacts of COVID-19 on rural populations in the U.S., Mueller et al. found extensive reports of the negative consequences from COVID-19 among rural populations including in unemployment rates, perceptions of the local economy, and impact on their overall lives and mental wellbeing (*60*). They reported a 9.74 percentage point increase in the 2020 unemployment rate in the rural communities included in their sample compared to the year before the pandemic (*60*). This contrasts with a national increase in the unemployment rate of only 7.40 percentage points. With increasing urbanization, rural populations represent a unique, and diverse community in the U.S., one particularly vulnerable to long-term social and economic consequences from COVID-19 (*61-63*). Taken together, this growing body of evidence suggests that both in urban and rural states, neighborhoods and communities that were structurally disadvantaged before the pandemic, and who experienced a high burden of COVID-19, may struggle to recover their social and economic wellbeing in the years post COVID-19.

The implications from our study for understanding the long-term social and economic fallout the U.S. is facing from the COVID-19 pandemic reveal the tip of an iceberg that warrants further investigation at the neighborhood-level. Structurally disadvantaged communities (e.g., low SES and/or communities of color) have experienced substantially greater population losses from COVID-19. A study of excess mortality conducted early on in the pandemic found that not only did racialized minority populations experience greater excess mortality compared to non-Hispanic White populations, but the inequity was even greater among working age adults (*64*). Non-Hispanic Black adults aged 35-44 years had a mortality rate ratio 9.0 times that of non-Hispanic Whites (*64*). A 2021 study found that racialized minority populations also experienced a greater number of COVID-19 hardships including food insufficiency, loss of income, unemployment, and housing instability and had a slower recovery from these hardships compared to their White counterparts.(21) A policy report from the Detroit Metro Area Communities Study (DMACs) found that the unemployment rate in Metro Detroit remained at 20% over the course of 2021, nearly twice the pre-pandemic rate of 10%.(*65*) Nationally, Black business owners and entrepreneurs were 30 times less likely than whites to receive federal COVID-19 relief funds (e.g., Paycheck Protection Program),(*66*) which will further magnify inequities in neighborhood resources.

Broadly, this work also suggests that that we need to move beyond a one-dimensional conceptualization of neighborhood poverty when trying to understand inequities in infectious disease, both the burden of illness and the social/economic consequences (*14, 67, 68*). Multiple forces converge to shape the health and wellbeing of a neighborhood and the risks associated with living in a given neighborhood—risks and benefits that are subsequently felt by neighborhood residents. These forces can include national and subnational policies, both historical and contemporary, that determine which groups of people live where (*69-71*). State policies trickle down to affect local policy in numerous ways, and are often dependent on the particular state context. As has been seen increasingly with COVID-19, political partisanship at multiple decision-making levels determines which mitigation strategies are put in place and how they are perceived and utilized by their targeted populations. These forces then interact with neighborhood-level social and economic indicators of disadvantage and affluence that collectively result in the contexts that put people at risk for infectious disease, and may or may not provide the resources to mitigate long-term health, social, and economic consequences (*50*). Thus, the prevailing assumption that neighborhood poverty is associated with increased infectious disease risk does not capture the heterogeneity in these associations brought on by the complex social forces that have shaped a particular neighborhood.

### Strengths & Limitations

We are among the first data collation efforts to attempt to collect spatially-referenced case data at the neighborhood-level for the entire U.S., and the only to link these spatially-referenced case data at the census tract or ZIP code level with novel features of the neighborhood built and social environment from the National Neighborhood Data Archive (NaNDA) repository. Our results have implications for both public health practice and policy and for how we understand the health of the U.S. population going forward. They also lay the groundwork for developing effective prevention and mitigation strategies to address continuing inequities in pandemic-related consequences. These data provide the foundation for future studies focused on the impacts of pandemic-related changes to the neighborhood environment and population health trends, and highlight the need to gather robust neighborhood-level disease, long-term sequelae, and environmental metrics for COVID-19 across the U.S. given the vast heterogeneity we observed by state.

While this study currently advances our understanding of how COVID-19 has differentially impacted neighborhoods across 21 states in the U.S., there are limitations that should be considered when interpreting these results and that may also guide future investigations. Given the emergency nature of the pandemic and the independence of state public health institutions from federal jurisdiction, each state set up surveillance systems differently, including defining different inclusion criteria and methods for counting cases, and in the selection of the types of health facilities surveilled (22-23). For example, if a state only included laboratory-confirmed cases, their case counts are likely an underestimate of the true case counts in a population, however, this was not standardized. Further, state case data often differed in geographic and temporal coverage, in part due to the rise in at home testing beginning from late 2021 onwards and other advances or policy changes, which made more nuanced comparisons by state difficult. While examining trends in the aggregate may mitigate some of the state-to-state differences, comparisons across states should still be made with caution; accounting for the decrease in case count accuracy over time is difficult even for the most robust surveillance teams.

Moreover, we only have data on incident infections. Thus, the case data could reflect repeat infections occurring in the same person. However, we can still detect neighborhoods that have had the highest COVID-19 burden. Additionally, we do not have information on co-morbid conditions occurring in tandem with COVID-19 infections which could be important drivers of COVID-19 infections and occur more frequently among structurally disadvantaged populations.(*72*) Future analyses should attempt to incorporate data on the prevalence of key co-morbid conditions into statistical models.

Importantly, as state case counts reflect not only the availability of testing facilities and infrastructure in place to report positive test results but access to such facilities as well, the current results may best be viewed as a proxy indicator of the true underlying trends in COVID-19 case data. In future studies, we plan to collate data on testing sites, test positivity rates, other indicators of the burden of COVID-19 (e.g., wastewater surveillance), and mitigation strategies (e.g., vaccination history) over time to create a more accurate indicator of COVID-19 burden.

Lastly, we believe that rurality may play a key role in the distribution of COVID-19 as well as how the neighborhood social environment relates to COVID-19 burden. Though we were not able to include an indicator of rurality in our models, there is a growing body of work showing a link between political partisanship and rurality. Therefore, it is possible that our political partisanship indicator may be capturing some of the rural/urban differences in these associations.(*73*)

Despite the limitations, these findings outline the necessary steps towards a more comprehensive data collation effort and possibly the foundation for a concerted surveillance system across the U.S. Compared to other U.S. spatially-referenced COVID-19 datasets like the Centers for Disease Control and Prevention COVID-19 Data Tracker or the Johns Hopkins University Coronavirus Resource Center, which are both limited to county-level data, harnessing a repository like CONEP will allow researchers and the public to examine more local level trends, acknowledging the heterogeneity that exists within counties.

As this is the first investigation in a series of studies stemming from CONEP, we will continue to build the CONEP repository to cover all 50 states and build in more contextual data at the state, county, and local levels. Also, it is critical to note that getting local-level, finer-scale spatially resolute data is difficult and should involve a deeper discussion of ethics, and calling on public health practitioners, who are often burdened with other real-time tasks, to collect such data may not always allow for the priority it should be given until systems are automated. Still, gathering these data is essential for monitoring health and social well-being in the U.S.

## Conclusion

The inequitable distribution of COVID-19 across neighborhoods in the U.S. has consequences both for the current health and economic wellbeing of the American population as well as for the future of population health in the U.S., and specifically population health inequities.(*74, 75*) Therefore, it is imperative that we interrogate patterns and trends at the local neighborhood-level. Our results document the complex ways in which the neighborhood social environment is related to COVID-19 burden and highlight the importance of the local-level for reducing disease risk. While the traditional paradigm in infectious disease research has long held that poverty increases infectious disease burden, our findings highlight extensive state-level variation, and that a one-size-fits-all approach will not address the unique patterns observed across states in the U.S. By leveraging fine-scale spatially-referenced case data, our findings enhance our understanding of the neighborhood social environment and COVID-19 burden and underscore the continued need for nationwide data for neighborhoods across all states.

## Methods

### The COVID Neighborhood Project

We launched CONEP in 2021 to address the need for fine-scale spatially resolute COVID-19 case data for the entire U.S. We contacted health departments in all 50 U.S. states. Initial contact consisted of calls, emails, and data portal requests asking for fine-scale COVID-19 case data. For some states, census tract or zip code data were publicly available. Others required a Freedom of Information Act (FOIA) request. Following request edits, resubmissions, and the eventual approval, we were sent data through a secure email. Data collation was done from June to August 2021.

### COVID-19 Data

We used COVID-19 cases at the zip-code level for sixteen states, and census tract-level for five states. Data were collected from state health departments throughout the summer and fall for both 2021 and 2022. Data were collected in a cumulative format. State COVID-19 cases were defined as they were by each respective state health department, most commonly as the sum of both laboratory confirmed and probable cases (1-3).

We collected both census tract and ZIP code level data. For those states with ZIP code level data, we used a cross-walk to merge ZIP codes into ZIP Code Tabulation Areas (ZCTA)s. ZIP codes are designated by the U.S. Postal Service and used to identify postal delivery routes. Therefore, they do not represent a spatial area. ZCTAs are generated by the U.S. Census Bureau and are generalized representations of ZIP codes. Methods used to create ZCTAs are detailed elsewhere (*76*).Of note, zip-code level data were not available in Florida past June 3rd, 2021, and Indiana past March 24th, 2022.

The study was approved by the Health and Behavioral Sciences Institutional Review Board at the University of Michigan. Data use agreements were done on a state-by-state basis (see Appendix).

### Estimates of social and physical context from the National Neighborhood Data Archive (NaNDA)

The primary goal of the current investigation was to report state-level trends in the distribution of COVID-19 burden at the neighborhood-level. However, the ultimate goal of CONEP is to examine how features of the social and physical environment have shaped COVID-19 burden throughout the pandemic, providing a roadmap for addressing the long-term consequences certain communities will face. As a first step, we estimated associations between state COVID-19 case counts and four indicators of the neighborhood social and/or physical environment: neighborhood affluence, neighborhood disadvantage, neighborhood population density, and county-level political partisanship.

We obtained measures of the neighborhood context from the National Neighborhood Data Archive (NaNDA), a publicly available repository of curated measures of social and physical environment context. We focused on three types of contextual measures hypothesized to be related to COVID-19 burden: neighborhood SES (disadvantage and affluence), neighborhood population density, and county-level political partisanship.

**Neighborhood disadvantage** is an analytically-derived index and is the mean of four variables collected as part of the American Community Survey: proportion of female-headed households with children, proportion of households receiving public assistance income or food stamps, proportion of families with income below the federal poverty level, and proportion of the population aged 16 years and older that are unemployed (*77*). Mean scores range from 0-100. Disadvantaged neighborhoods tend to have fewer resources (e.g., healthy food stores, well-maintained parks, good schools, quality medical care) (*18, 78*) and are often vulnerable to disinvestment and environmental hazards (*79*).

**Neighborhood affluence** is the mean of three variables from the American Community Survey: percent of household with income greater than $75K per year, percent of the population over the age of 16 employed in professional or managerial occupations, and percent of the population with a Bachelor’s Degree or higher (*77*). Mean scores range from 0-100. Higher values indicate a more affluent neighborhood. Affluent neighborhoods are likely to attract a set of institutions (e.g., food stores, places to exercise, well-maintained buildings and parks) that foster a set of norms (e.g., an emphasis on exercise and healthy diets) conducive to good health.(*80*) Both from a theoretical and analytical standpoint, neighborhood affluence is distinct from neighborhood disadvantage. (*77*) Neighborhood affluence is associated with higher levels of social control and leverage over local institutions that can foster social environments that facilitate health.(*14*)

**Neighborhood population density** is an indicator of the number of persons per square mile in the census tract and/or ZCTA (*77*). We included neighborhood population density to proxy the potential for exposure to the SARS-COV-2 pathogen within a given neighborhood with the hypothesis that those neighborhoods with higher population density would have higher COVID-19 case counts.

We defined **county-level political partisanship** with a continuous measure that indicates the percent of votes cast for Republican candidates in 2018 and the six years before (*81*). We linked ZCTAs and census tracts to the respective counties in which they resided. For ZCTAs that spanned two counties, we chose the county that had the greatest proportion of the ZCTA.

### Statistical Analyses

We first calculated the cumulative COVID-19 case count per 100,000 population for each state. We then calculated the median case count per 100,000 and interquartile range (IQR) for ZCTAs or census tracts within states cumulatively for the entire time period for which the state reported data.

We then constructed a series of Poisson regression models to estimate the association between neighborhood characteristics and the incidence rate ratio (IRR) of COVID-19 cases for neighborhoods within each state. The total population of the neighborhood served as the offset term in the models. We estimated models examining two major exposures: neighborhood disadvantage and neighborhood affluence. In model 1, we controlled for neighborhood population density and in model 2 we added an additional control variable for county-level political partisanship. We used generalized estimating equations with robust standard errors to account for clustering of census tracts and ZCTAs in counties.

All statistical analyses were carried out in Stata/mp 17.0.

## Statements

### COVID-19 Data Health Department Acknowledgements

Data for Arizona were obtained through public download from the Arizona Department of Health Services (ADHS). ADHS is not responsible for the author’s analysis, opinions, and conclusions contained in this document.

Data for Delaware were obtained through public download from the Delaware Department of Health and Social Services (DHSS). DHSS is not responsible for the author’s analysis, opinions, and conclusions contained in this document.

Data for Florida were obtained through public download from the Florida Department of Health (FDOH). FDOH is not responsible for the author’s analysis, opinions, and conclusions contained in this document.

Data for Illinois were obtained through public download from the Illinois Department of Public Health (IDPH). IDPH is not responsible for the author’s analysis, opinions, and conclusions contained in this document.

Data for Indiana were obtained through public download from the Indiana Department of Health (IDOH). IDOH is not responsible for the author’s analysis, opinions, and conclusions contained in this document.

Data for Louisiana were obtained through public download from the Louisiana Department of Health (LDH). LDH is not responsible for the author’s analysis, opinions, and conclusions contained in this document.

Data for Maine were obtained through public download from the Maine Department of Health and Human Services (Maine DHHS). Maine DHHS is not responsible for the author’s analysis, opinions, and conclusions contained in this document.

Data for Maryland were obtained through public download from the Maryland Department of Health (MDH). MDH is not responsible for the author’s analysis, opinions, and conclusions contained in this document.

Data for Minnesota were obtained through a data practices request to the Minnesota Department of Health (MDH). MDH is not responsible for the author’s analysis, opinions, and conclusions contained in this document.

Data for Nevada were obtained through a public records request to the Nevada Department of Health and Human Services (Nevada DHHS), and the assistance of Kavita Batra. Nevada DHHS is not responsible for the author’s analysis, opinions, and conclusions contained in this document.

Data for New Mexico were obtained through a public records request from the New Mexico Department of Health (NMDOH). NMDOH is not responsible for the author’s analysis, opinions, and conclusions contained in this document.

Data for New York were obtained through public download from the New York State Department of Health (NYSDOH). NYSDOH is not responsible for the author’s analysis, opinions, and conclusions contained in this document.

Data for North Carolina were obtained through public download from the North Carolina Department of Health and Human Services (NCDHHS). NCDHHS is not responsible for the author’s analysis, opinions, and conclusions contained in this document.

Data for Ohio were obtained through public download from the Ohio Department of Health (ODH). ODH is not responsible for the author’s analysis, opinions, and conclusions contained in this document.

Data for Oklahoma were obtained through public download from the Oklahoma State Department of Health (OSDH). OSDH is not responsible for the author’s analysis, opinions, and conclusions contained in this document.

Data for Oregon were obtained through public download from the Oregon Department of Human Services (ODHS). ODHS is not responsible for the author’s analysis, opinions, and conclusions contained in this document.

Data for Pennsylvania were obtained through public download from the Pennsylvania Department of Health (PDH). PDH is not responsible for the author’s analysis, opinions, and conclusions contained in this document.

Data for Rhode Island were obtained through a request to the Center for Health Data and Analytics, COVID-19 Quant Team, Rhode Island Department of Health (RIDOH). RIDOH is not responsible for the author’s analysis, opinions, and conclusions contained in this document.

Data for Vermont were obtained through a public records request to the Vermont Department of Health (VDH), and the assistance of Lucy Lincoln. VDH is not responsible for the author’s analysis, opinions, and conclusions contained in this document.

Data for Virginia were obtained through public download from the Virginia Department of Health (VDH). VDH is not responsible for the author’s analysis, opinions, and conclusions contained in this document.

Data for Wisconsin were obtained through public download from the Wisconsin Department of Health Services (Wisconsin DHS). Wisconsin DHS is not responsible for the author’s analysis, opinions, and conclusions contained in this document.

## Funding Acknowledgements

This study was partially funded by the U.S. National Institutes of Health, National Institute on Aging Network on Life Course Dynamics and Disparities in the 21^st^ Century R24 AG045061 (PI Noppert). Funding support was also provided by the National Institutes of Health, National Institute of Nursing Research and National Institute of Minority Health and Health Disparities U01 NR020556 (MPI Noppert and Clarke). KAD received support for this work from the National Institutes of Health, National Institute on Aging R00 AG066846 and JTK received support for this work from the National Institutes of Health, National Institute on Aging U24 AG076462.

## Data Availability

Details regarding data availability are described separately for each state health department.

## Acknowledgements

The authors are deeply grateful to the health departments who shared their data with us. Additionally, this work has been done as part of the Social Environmental and Equity in Infectious Disease Lab—a interdisciplinary collaborative between the University of Michigan and Johns Hopkins University.

