## Supplementary File for "State Variation in Neighborhood COVID-19 Burden: Findings from the COVID Neighborhood Project"

**Supplementary Material**

**Figures**

**Figure S1.** Map showing the states currently included in the COVID Neighborhood Project (CONEP). The case counts per 100,000 population categorized into deciles for all 21 states are depicted. The case counts refer to cumulative case counts for the time period. Color gradations refer to higher deciles of case counts.

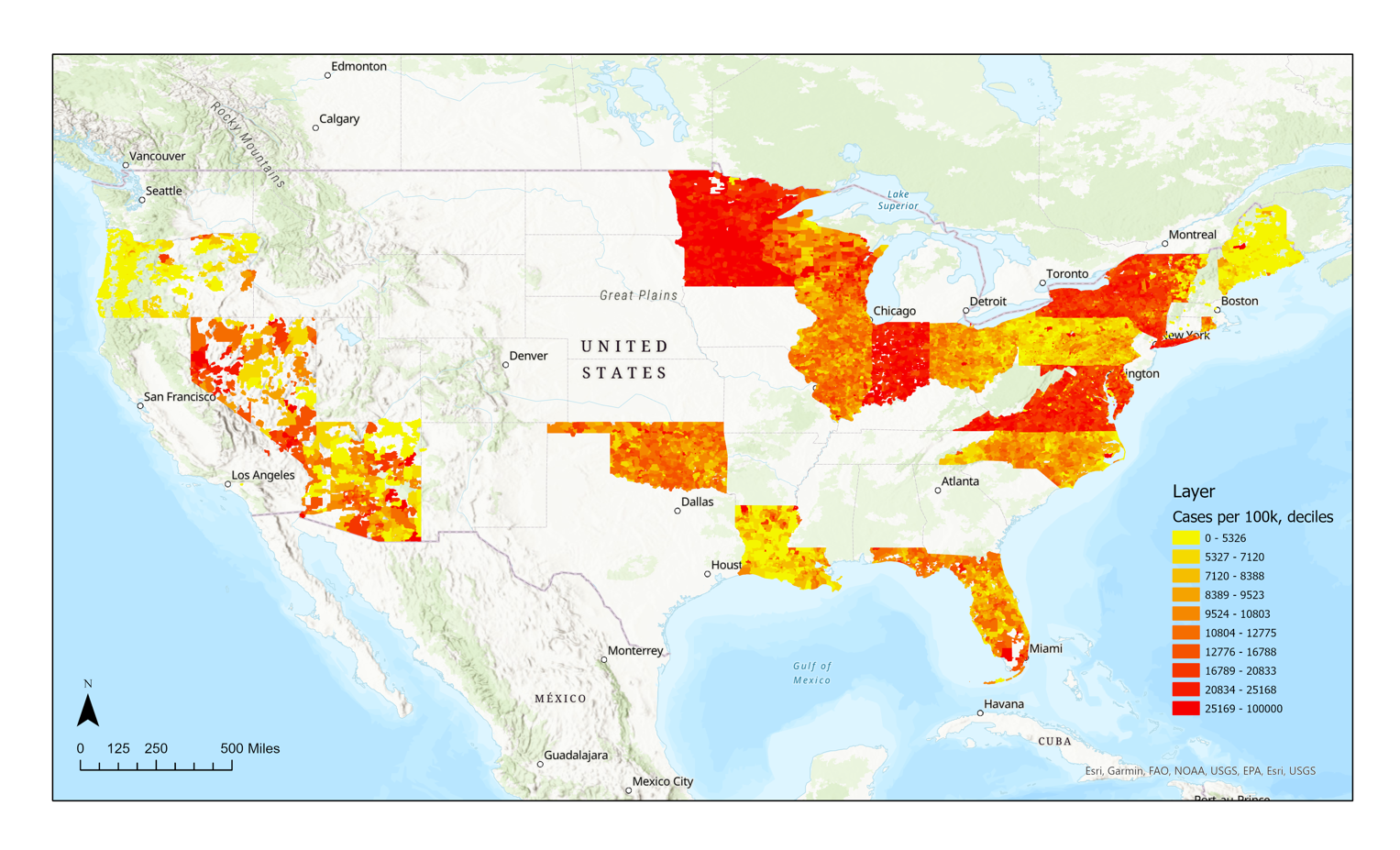

**Figure S2.** Map showing the case counts per 100,000 persons for Oregon, Nevada, and Arizona. The case counts per 100,000 population categorized into deciles for all 21 states are depicted. The case counts refer to cumulative case counts for the time period. Color gradations refer to higher deciles of case counts.

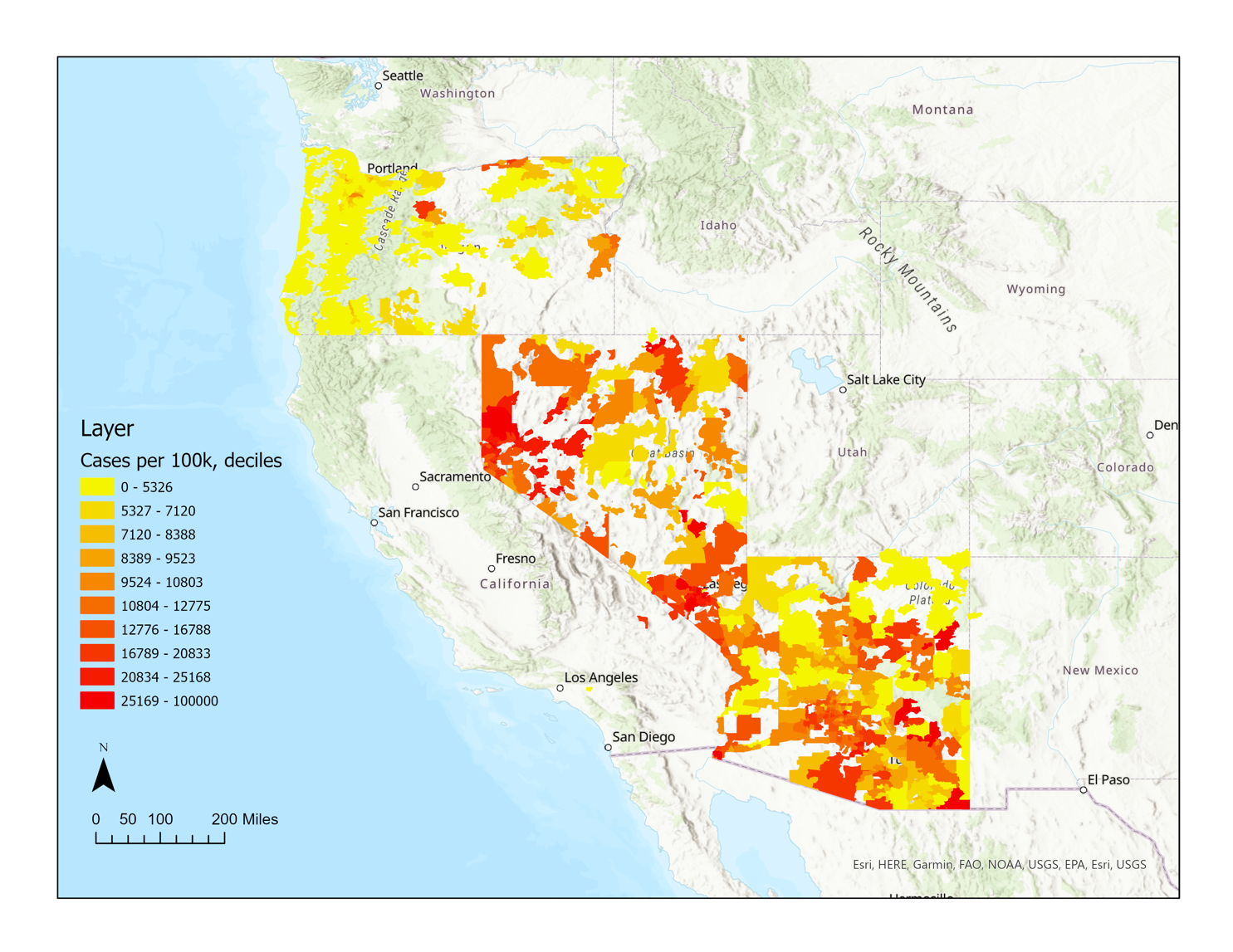

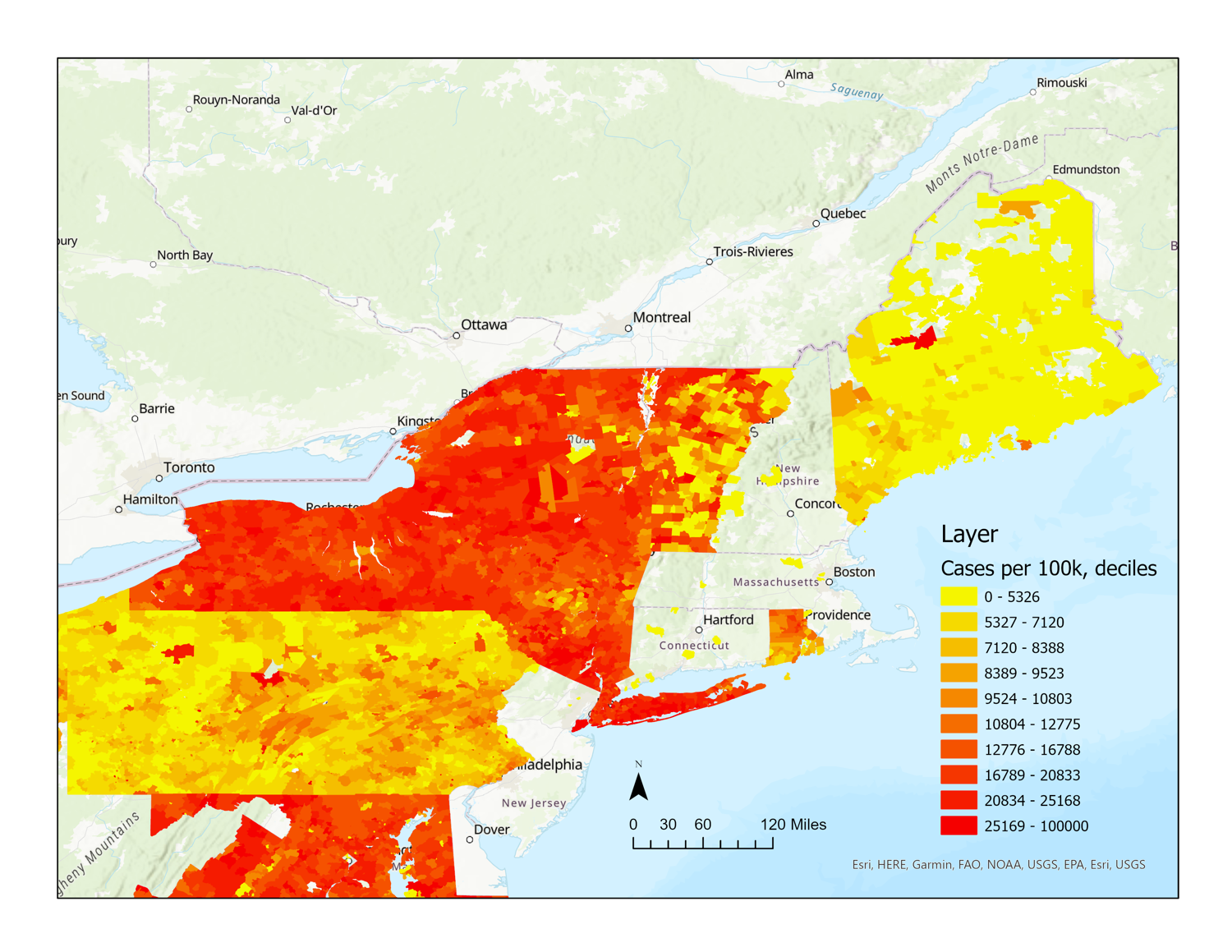
**Figure S3.** Map showing the case counts per 100,000 persons for Maine, New York, Rhode Island, and Pennsylvania. The case counts per 100,000 population categorized into deciles for all 21 states are depicted. The case counts refer to cumulative case counts for the time period. Color gradations refer to higher deciles of case counts.

**Tables**

**Table S1.** Number of spatial units in each state missing data.

| **State** | **Spatial**  **Resolution** | **Number of Spatial Units Missing Data/ Total Spatial Units in the State**** |
| --- | --- | --- |
| **Northeast** | | |
| Delaware | Census tract | 38/214 |
| Maine | ZCTA | 38/427 |
| Maryland | ZCTA | 21/464 |
| New York | ZCTA | 0/1,753 |
| Pennsylvania | ZCTA | 50/1,783 |
| Rhode Island | Census tract | 0/77 |
| Vermont | ZCTA | 0/254 |
| **Southwest** | | |
| Arizona | ZCTA | 48/397 |
| New Mexico | Census tract | 4/603 |
| Oklahoma | ZCTA | 20/646 |
| **West** | | |
| Nevada | ZCTA | 1/171 |
| Oregon | ZCTA | 143/415 |
| **Southeast** | | |
| Florida | ZCTA | 36/976 |
| Louisiana | Census tract | 10/1,127 |
| North Carolina | ZCTA | 41/801 |
| Virginia | ZCTA | 0/891 |
| **Midwest** | | |
| Illinois | ZCTA | 31/1,381 |
| Indiana | ZCTA | 251/771 |
| Minnesota | ZCTA | 8/880 |
| Ohio | ZCTA | 52/1,188 |
| Wisconsin | Census tract | 0/1,392 |
| **Indicates the total number of spatial units with population that are missing values | | |

**Table S2.** Results of the regression analyses estimating the cumulative incidence rate ratio of neighborhood COVID-19 burden by quartiles of neighborhood disadvantage. Model 1 controls for neighborhood population density and model 2 includes an additional control for county-level political partisanship.

2a.

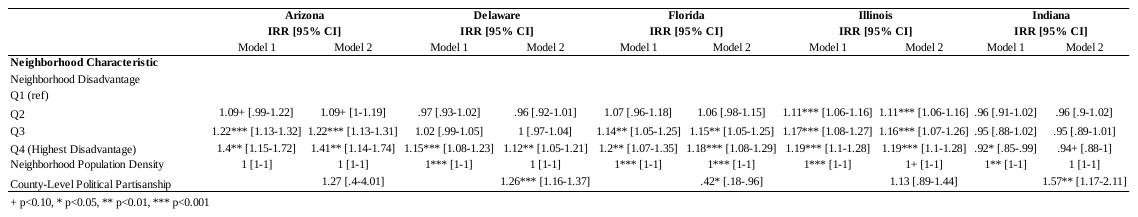

2b.

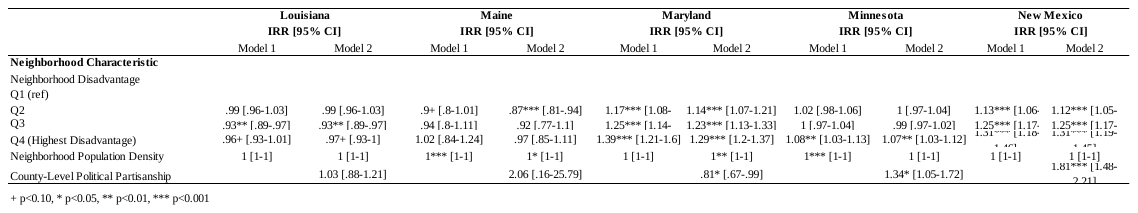

2c.

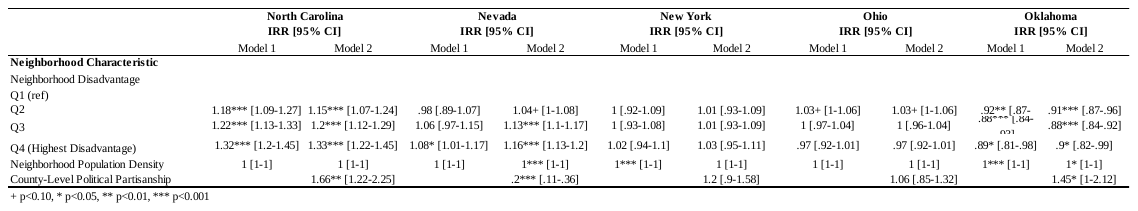

2d.

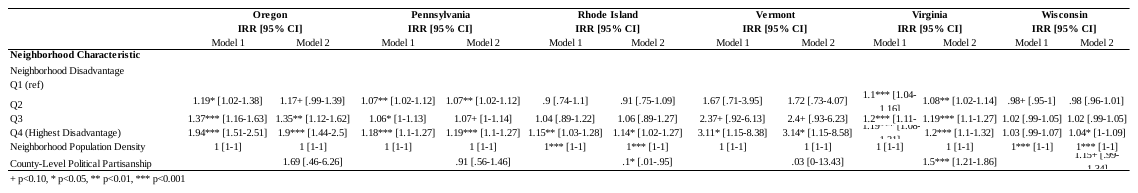

**Table S3.** Results of the regression analyses estimating the cumulative incidence rate ratio of neighborhood COVID-19 burden by quartiles of neighborhood affluence. Model 1 controls for neighborhood population density and model 2 includes an additional control for county-level political partisanship.

3a.

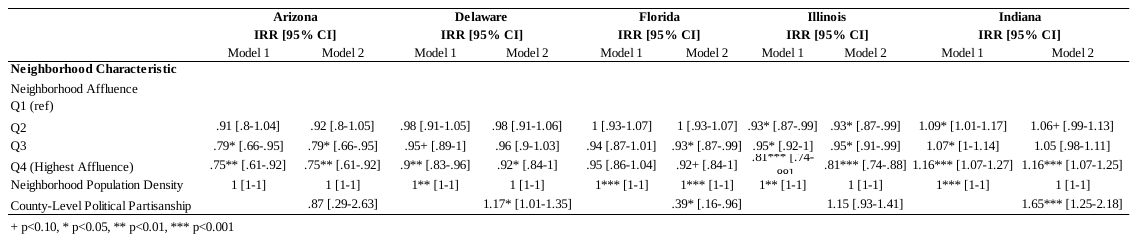

3b.

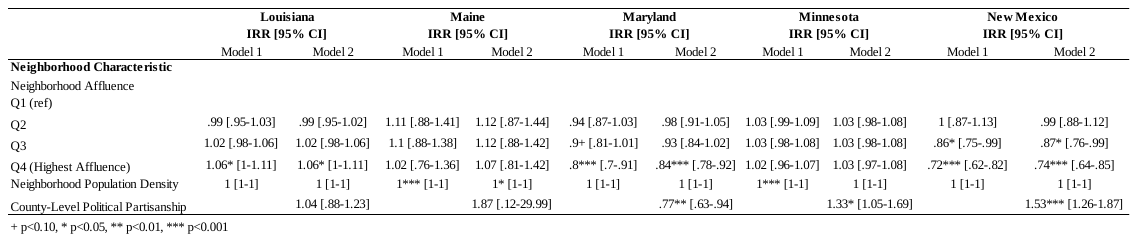

3c.

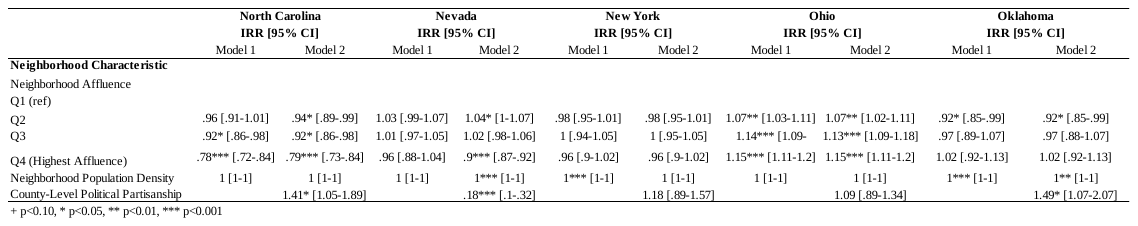

3d.

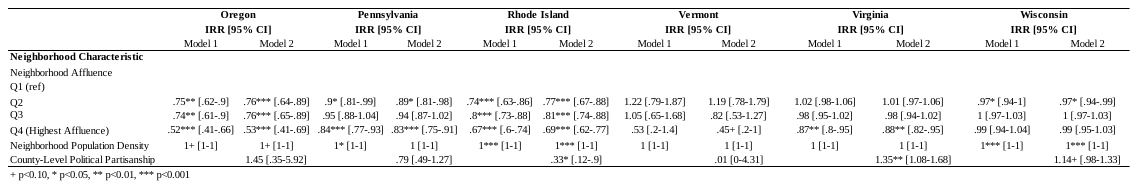
